# Covid-19 automated diagnosis and risk assessment through Metabolomics and Machine-Learning

**DOI:** 10.1101/2020.07.24.20161828

**Authors:** Jeany Delafiori, Luiz Claudio Navarro, Rinaldo Focaccia Siciliano, Gisely Cardoso de Melo, Estela Natacha Brandt Busanello, José Carlos Nicolau, Geovana Manzan Sales, Arthur Noin de Oliveira, Fernando Fonseca Almeida Val, Diogo Noin de Oliveira, Adriana Eguti, Luiz Augusto dos Santos, Talia Falcão Dalçóquio, Adriadne Justi Bertolin, João Carlos Cardoso Alonso, Rebeca Linhares Abreu-Netto, Rocio Salsoso, Djane Baía-da-Silva, Vanderson Souza Sampaio, Carla Cristina Judice, Fabio Trindade Maranhão Costa, Nelson Durán, Mauricio Wesley Perroud, Ester Cerdeira Sabino, Marcus Vinicius Guimarães Lacerda, Leonardo Oliveira Reis, Wagner José Fávaro, Wuelton Marcelo Monteiro, Anderson Rezende Rocha, Rodrigo Ramos Catharino

**Author notes:** Corresponding author: RRC –; ARR –. The authors equally contributed to the work (Jeany Delafiori, Luiz Claudio Navarro, Rinaldo Focaccia Siciliano).

## Abstract

COVID-19 is still placing a heavy health and financial burden worldwide. Impairments in patient screening and risk management play a fundamental role on how governments and authorities are directing resources, planning reopening, as well as sanitary countermeasures, especially in regions where poverty is a major component in the equation. An efficient diagnostic method must be highly accurate, while having a cost-effective profile.

We combined a machine learning-based algorithm with instrumental analysis using mass spectrometry to create an expeditious platform that discriminate COVID-19 in plasma samples within minutes, while also providing tools for risk assessment, to assist healthcare professionals in patient management and decision-making. A cross-sectional study with 728 patients (369 confirmed COVID-19 and 359 controls) was enrolled from three Brazilian epicentres (São Paulo capital, São Paulo countryside and Manaus) in the months of April, May, June and July 2020.

We were able to elect and identify 21 molecules that are related to the disease’s pathophysiology and 26 features to patient’s health-related outcomes. With specificity >97% and sensitivity >83% from blinded data, this screening approach is understood as a tool with great potential for real-world application.

## INTRODUCTION

Coronaviruses (CoVs) are enveloped, single-stranded positive RNA viruses from the *Coronaviridae* family (1). The recent pandemic, caused by a newly discovered strand of coronavirus, SARS-CoV-2, was denominated COVID-19 (2), a disease that disseminated fast and is responsible for hundreds of thousands of deaths worldwide. Measures to control disease spread have led most countries to adopt social distancing and population screening (3). Given its global economic, sanitary and social impact, thousands of new studies aiming to understand viral pathology and targets for virus dissemination control are being conducted, which directly impact in strategies to provide treatments, vaccines, screening tests and patient prognosis.

Special efforts have been directed towards the development of alternatives for mass testing with population-wide capabilities. Currently, available tests are based on the direct detection of

SARS-CoV-2 virus through antigens or RNA amplification (RT-PCR), serological tests to evaluate patient immunity, and the combination of RT-PCR and chest CT (computed-tomography). COVID-19 testing urgency comprises the need for medical decision-making tools for patient’s risk stratification and management, which is poorly achieved by standard methodologies. Even though the basis for these procedures are well-documented in the literature, there are increased concerns about test’s sensitivity and specificity achieved on the field, time and costs associated with procedures, reagents and trained personnel availability, and the testing window (4-6).

Difficulties for an accurate diagnosis of SARS-CoV-2 and patient’s risk categorization are consequences of COVID-19 complexity. SARS-CoV-2 infection pathophysiology reflects in a broad spectrum of patient symptoms, ranging from mild flu-like manifestations, such as fever, cough, and fatigue, to life-threatening acute respiratory distress syndrome (ARDS), vascular dysfunction, and sepsis (2, 7). In an effort to eliminate the pathogen, the body response to SARS-CoV-2 severe pulmonary infection involves the reduction of natural killer (NK) cells, increased pro-inflammatory cytokines (IL-6, IFN-γ, TNFα and others) and lung infiltration, especially by macrophages and monocytes (2, 7, 8), possibly resulting in tissue damage and organ injury (8, 9).

Furthermore, changes in lipid homeostasis, a common characteristic of viral infections, have been associated with SARS-CoV-2 pathology (9-11). In lipidomic and metabolomic profiling of plasma samples, Song et. al. (2020) suggested that exosomes enriched with monosialodihexosyl ganglioside (GM3) are associated with the severity of COVID-19. In the same study, the decrease of circulating acyl-carnitines indicates disturbance in oxidative stress and cellular energy support (9). Moreover, Fan et al. (2020) proposed the relationship between progressive decrease in serum low-density lipoprotein (LDL) and cholesterol within deceased patients (10). Moreover, individual susceptibility to COVID-19 symptoms are not fully understood, thereby hampering any potential outcome prediction.

Panels of biomarkers that translate disease pathophysiology and contribute to SARS-CoV-2 detection may be proposed through “omics” techniques (9, 11, 12). The current trend in associating artificial intelligence-explained algorithms and “omics” techniques has yielded platforms involving machine learning (ML) to analyze mass spectrometry (MS) data, aiming at biomarker identification of diseases, including COVID-19 severity assessment (11, 13). However, applying traditional untargeted mass spectrometry for diagnostic purposes is laborious, since it requires further method development and validation steps (13, 14).

Considering that the testing tool for COVID-19 introduced in this contribution is based on metabolites from actual patients, it may be considered a new approach for SARS-CoV-2 screening. The proposed end-to-end mass spectrometry and machine learning combination aims at predictively identifying and modeling putative biomarkers for COVID-19 identification and risk assessment. This is critical for effective implementation on a real-world setting, adding robustness to the model in spite of variations in the input data; issues due to noise and minor different variations in acquisition conditions will, therefore, not play a major interference in the final output. Therefore, using the potential of MS-ML techniques in COVID-19 fighting (15), we enrolled a cohort of 728 individuals for the development of this independent platform that simultaneously functions as an automated screening test using plasma samples with high specificity and sensitivity, and provides metabolic information related to the presence and severity risk for the disease.

## METHODS

### Study design and patient recruitment

Participants were recruited from selected sites with proven expertise in research and high volume of patients with COVID-19 to increase data variability: Central Institute of the Clinical Hospitals, University of São Paulo Medical School (localized in São Paulo, capital of the São Paulo State), Sumaré State Hospital (localized in the state of São Paulo inland), and Hospital Delphina Rinaldi Abdel Aziz (localized in Manaus, capital of Amazonas State localized in the North of the country). The study was conducted according to principles expressed in Declaration of Helsinki and approved by local Ethics Committees (CAAE 32077020.6.0000.0005, CAAE 31049320.7.1001.5404 and CAAE 30299620.7.0000.0068). Inclusion criteria for COVID-19 group (CV) were adult patients with one or more clinical symptoms of SARS-CoV-2 infection in the last seven days (fever, dry cough, malaise and/or dyspnea) and positive SARS-CoV-2 RT-PCR in nasopharyngeal samples, following local hospital testing protocols based on Charité protocol and WHO recommendations (16). A control group (CT) was formed by symptomatic RT-PCR-negative participants (SN) with SARS-CoV-2 discarded by clinical and tomographic picture, and non-infected controls (AS).

In this study, 728 participants were included, classified according to symptoms, RT-PCR testing results and respective risk (***Figure 1a***). CV was composed of 487 plasma samples from 369 symptomatic SARS-CoV-2 confirmed cases upon hospital arrival, and 118 samples representing a second collection from hospitalized patients (median 11 days, SD 3.8) that recovered (R) or deceased (D). The high-risk group (HRSP) comprised patients with moderate and severe symptoms that required hospitalization (n = 197) and the low-risk (LRSP) category (n = 172) contained those with mild symptoms redirected to home care. Gender, age, and fasting restrictions were not applied, to simulate real-world conditions and to provide results with no patient bias. CT group was formed by 29 SN and 330 AS, totaling 359 individuals ***Table S1*** (supplementary material) shows detailed demographic information and participant breakdown.

**Figure 1.**
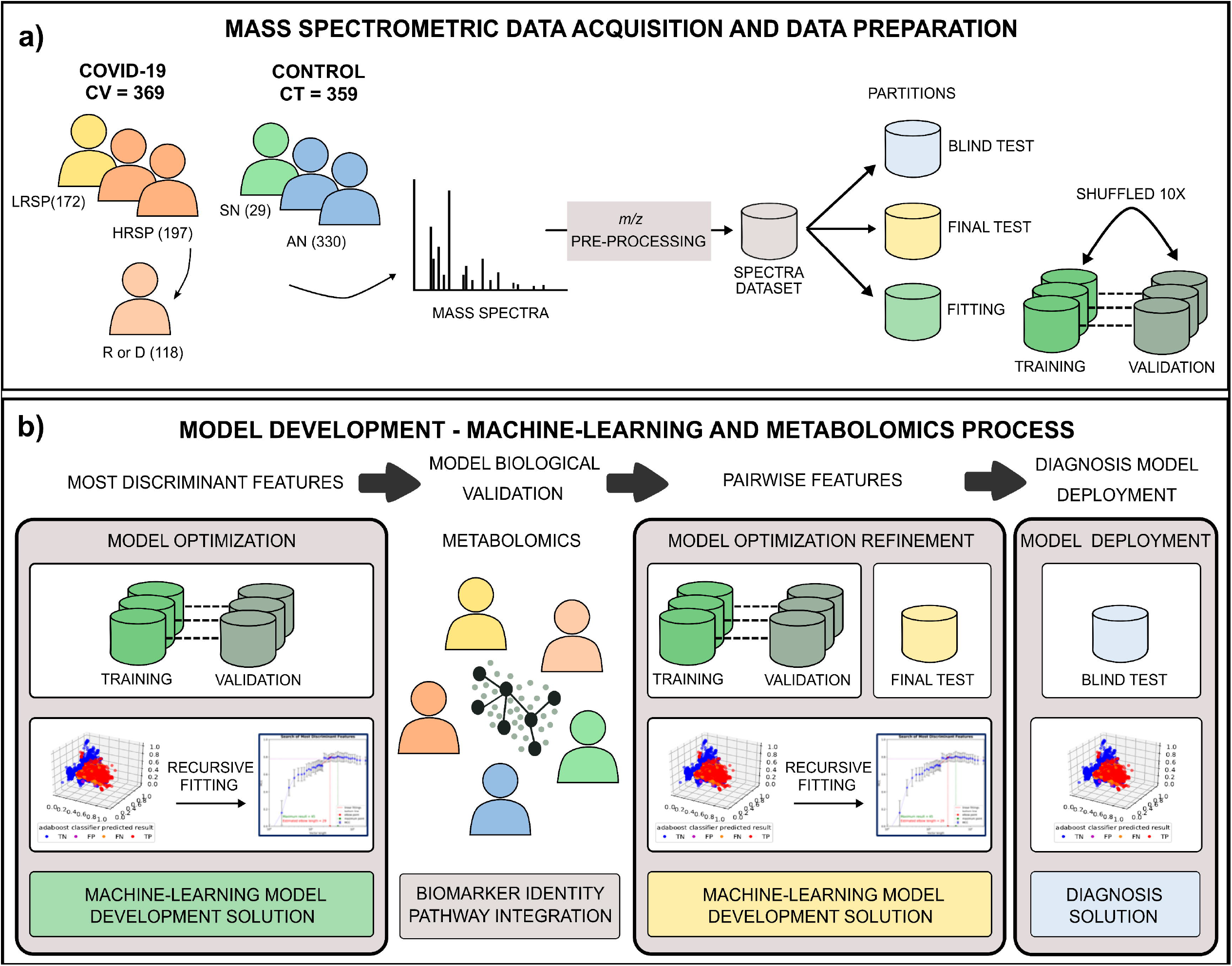
End to end process for putative biomarkers determination and diagnosis test generation. a) Based on clinical symptoms and diagnosis results, subjects were grouped in low-risk symptomatic positive (LRSP), high-risk symptomatic positive (HRSP), recovered (R) or deceased (D), symptomatic negative (SN) and asymptomatic negative (AN). Samples were prepared injected in a high-resolution mass spectrometry (HR-MS) equipment for data acquisition and datasets generated for data analysis according to the partitions; b) Sequential steps of machine learning data analysis and metabolomics biomarkers determination were followed for diagnosis model generation and deployment.

### Mass spectrometry sample preparation

Plasma samples from peripheral venous blood were frozen at -80°C until analysis. A 20-µL aliquot of each participant plasma was diluted in 200 µL of tetrahydrofuran, followed by homogenization for 30 seconds at room temperature. Thus, 780 µL of methanol was added followed by a second homogenization for 30 seconds and centrifugation for 5 min, 3400 *x* rpm at 4°C. An aliquot of 5 µL of the supernatant was diluted in 495 µL of methanol and positively ionized by the addition of formic acid (0·1% final concentration) prior to direct infusion in a high-resolution mass spectrometer.

### Mass spectrometry analysis and biomarker elucidation

Samples from CT and CV groups were randomized for data acquisition intra- and inter-daily. Samples were directly infused in a HESI-Q-Orbitrap®-MS (Thermo Scientific, Bremen, Germany) and scanned with 140,000 FWHM of mass resolution on positive ion mode. MS parameters were set as follows: *m/z* range 150-1,700, 10 mass spectral acquisition per sample, sheath gas flow rate five units, capillary temperature 320°C, aux gas heater temperature 33°C, spray voltage 3·70 kV, automatic gain control (AGC) at 1 × 10^6^, S-lens RF level 50, and injection time < 2 ms. After machine leaning modeling, the presence of each discriminant *m/z* determined by the algorithm was confirmed in mass spectra using Xcalibur 3.0 software (Thermo, Bremen, Germany). Molecule identification was proposed using METLIN (Scripps Center for Metabolomics, https://metlin.scripps.edu), HMDB (Human Metabolome Database, http://www.hmdb.ca/) and LIPIDMAPS (Lipidomics Gateway, https://lipidmaps.org) databases and literature search with mass accuracy ≤ 5 ppm.

Biomarker pathway analysis and meaning were attributed based on Kegg database (Kyoto Encyclopedia of Genes and Genomes, https://www.genome.jp/kegg/) information and scientific literature.

### Machine learning data analysis

The MS-ML platform presented in this study for COVID-19 automated diagnosis and risk determination consists of two primary data analysis phases. The first phase comprises developing a machine-learning model (ML) using a classification algorithm over MS data to determine potential *m/z* biomarkers for diagnosis and risk determination. The second phase entails a prediction model for diagnosing and determining a high-risk versus low-risk program, which will be used for individuals screening in the field.

Data processing is divided into the sequential steps described in ***Figure 1***. First, mass spectrometric data are pre-processed for ion annotation (intensity, width, resolution, and *m/z* values), alignment, normalization, and denoising. Three different partitions of resulting data are segregated according to the best practices of machine learning, consisting of a fitting partition (training and validation, shuffled in all ten rounds of ML experiments), test partition, and blind test. The final classification results are reported using the blind partition (see process in ***Figure 1a***).

The most discriminant features are determined using the ML algorithms (ADA Tree Boosting (ADA), Gradient Tree Boosting (GDB), Random Forest (RF), and Extreme Random Forest (XRF), which are based on decision trees. In addition, we also explored the Partial Least Squares (PLS) method, which is a linear space transformation (17-19)), in which a recursive fitting is applied to training and validation data (see ***Figure 1b***), with the annotation of averaging and computing the related standard deviation of selected performance metrics.

In all experiments, we adopted the performance metrics defined in ***Table S2*** (supplementary material) for each round of validation (optimized through accuracy, F1score, MCC). After the observation of performance metrics versus ranked features length, discriminant *m/z* features are evaluated through ΔJ importance (see ***Table S2*** and ***Figure 2a***) and selected for metabolomics biomarkers identification (see section *Mass Spectrometry Analysis and Biomarker Elucidation*). The marker importance is given by a cumulative distribution function (CDF) analysis: for a specific m/z, a CDF of the feature values for the negative samples (CT group) is compared with the CDF of positive samples (CV group) used in the fitting partition.

**Figure 2.**
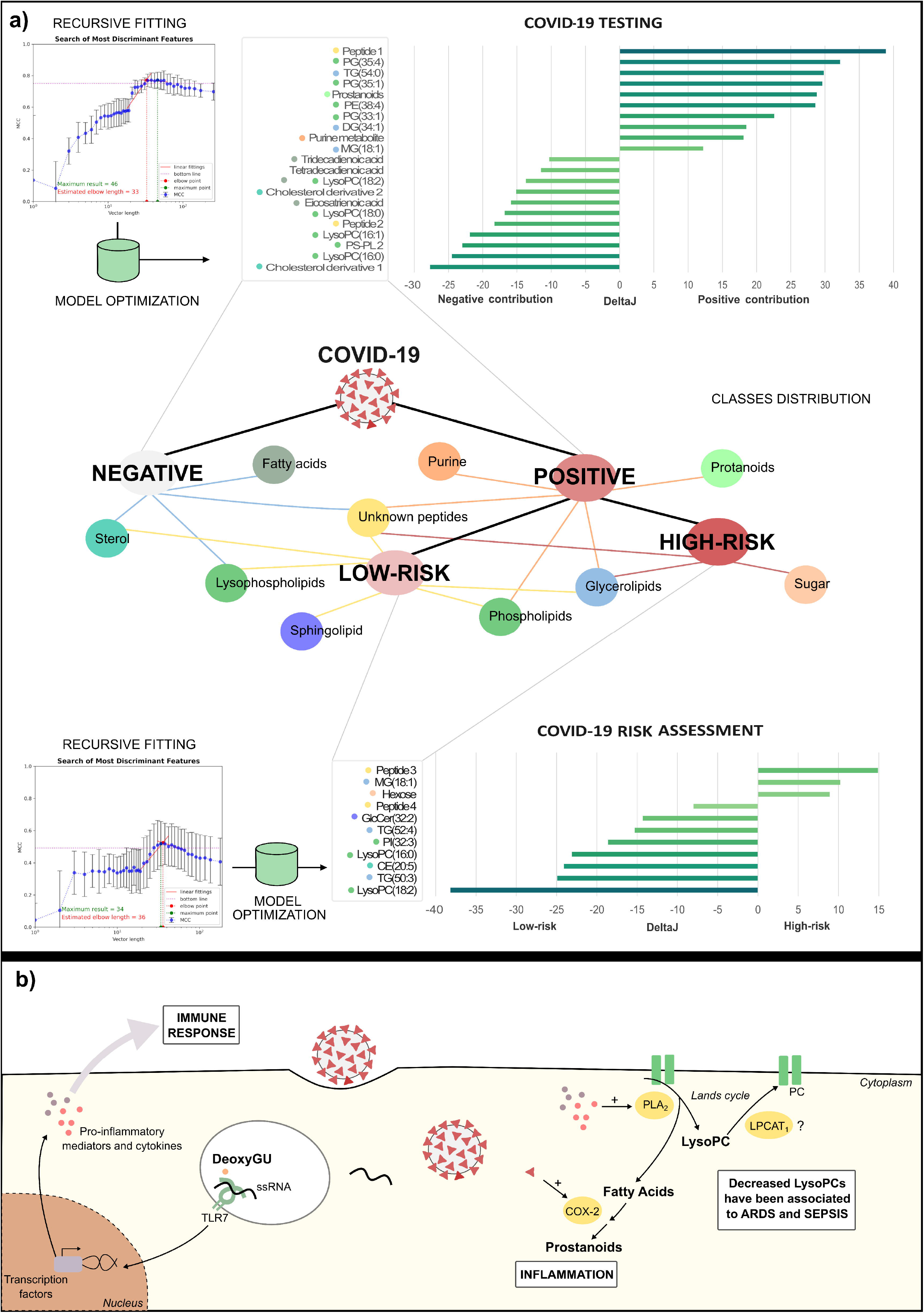
Putative biomarkers elucidation and related class/ pathways. a) Recursive fitting of mass spectra data followed by model optimization processes allowed the determination of putative biomarkers ranked by DeltaJ importance and group contribution. b) Proposed role of identified biomarkers in COVID-19 pathophysiology. Abbreviations: ARDS – acute respiratory distress syndrome, COX-2 – cyclooxygenase-2, DeoxyGU, deoxyguanosine, LPCAT1 - lysophosphatidylcholine acyltransferase 1, LysoPC – lysophosphatidylcholine, PC – phosphatidylcholine, PLA_2_ – Phospholipase A_2_

The CDF comparison uses first the Kolmogorov-Smirnov (KS-test) two samples equality hypothesis test to determine whether those distributions are different (failed on equality hypothesis). Then the ⍰J metric defined in ***Table S2*** is used to determine if the features contribute negatively ⍰J < 0, which means the negative samples (CT) have a higher probability of presenting higher values over the median of negatives, or positively ⍰J < 0, which means that the positive samples (CV) have a higher probability of presenting higher values over the median of positives. Features are discarded if CDFs are equal according to KS-test or ⍰J = 0. The selected biomarkers undergo a second round of training and validation with the algorithms mentioned above with the development software (***Figure 1b***, see testing results in ***Tables 2*** and ***3***). As putative biomarkers are validated through the development process, they are submitted to the second phase of the machine-learning process targeting the final model to deliver an applied untargeted metabolomics diagnosis software. In this phase, a pairwise model is created (***Figure 1b***), where the relationship between the putative biomarkers are used instead of their intensity (or relative abundance) provided in each spectrum.

## RESULTS

### COVID-19 testing through MS-ML platform: modeling and performance

The full dataset resulting from the spectrometer acquisition has 846 biological samples, with ten replicates each, on average. ***Table 1*** shows the data preparation for the fitting process (shuffled in 10 rounds of training and validation), and testing.

**Table 1.** Dataset subdivisions for model fitting (training and validation), testing and blind test for COVID-19 diagnosis and risk assessment.

| Model | COVID-19 diagnosis (n= 846) |  |  | COVID-19 risk assessment |  |  |
| --- | --- | --- | --- | --- | --- | --- |
| Class | Positive | Negative | Subtotal | High-risk | Low-risk | Subtotal |
| Fitting - Training | 219 | 195 | 414 on avg.<br>(49%) | 96 | 92 | 188 on avg.<br>(51%) |
| Fitting - Validation | 89 | 79 | 168 on avg.<br>(20%) | 39 | 37 | 76 on avg.<br>(21%) |
| Testing | 49 | 44 | 93 (11%) | 23 | 22 | 45 (12%) |
| Blind Test | 130 | 41 | 171 (20%) | 39 | 21 | 60 (16%) |

In this study, we employed a novel sequential processing of metabolomics data with Machine-Learning algorithms for building a model divided into two phases. First, a predictive modeling for putative biomarker identification. Then, a combination of biomarker features into relative pairs, composing the predictive model used by the diagnosis and risk assessment in the field (recursive fitting shown in ***Figure 1b***).

The analysis for diagnosis was performed with the full dataset, while the risk assessment relied on 369 COVID-19 positive subjects, as this is a second-stage analysis. Out of the COVID-19 positive subjects, 197 achieved local clinical criteria for hospitalization while the remaining 172 individuals were forwarded to homecare. ***Tables 2*** and ***3*** show results for the pairwise features for COVID-19 automated diagnosis and risk assessment classifiers, respectively. The best results were obtained with Gradient Tree Boosting (GDB): COVID-19 automated diagnosis with 97.6% of specificity and 83.8% of sensitivity, and risk assessment with 76.2% of specificity and 87.2% of sensitivity, both in the blind test.

**Table 2.** Performance metrics for diagnostics model using pairwise features on the 10 validation tests with 6 different classifier algorithms for Covid-19 positive/negative diagnosis, final development testing and deployed software blind test. Numbers correspond to individual’s classification average and standard deviations in parenthesis.

| Model | COVID-19 diagnosis development validation |  |  |  |  |  | Test | Blind Test |
| --- | --- | --- | --- | --- | --- | --- | --- | --- |
| Algorithms | ADA | GDB | RF | XRF | PLS | SVM | GDB | GDB |
| Vector length | 39 | 39 | 39 | 39 | 39 | 39 | 39 | 39 |
| # of Estimators | 260 (3) | 260 (3) | 260 (3) | 260 (3) | NA | NA | 256 | 256 |
| TN | 73 (3) | 72 (3) | 70 (3) | 69 (3) | 67 (4) | 71 (3) | 42 | 41 |
| FP | 7 (2) | 7 (2) | 9 (3) | 11 (3) | 12 (3) | 8 (2) | 2 | 1 |
| FN | 5 (2) | 4 (1) | 10 (4) | 7 (2) | 7 (2) | 9 (2) | 3 | 21 |
| TP | 84 (4) | 85 (3) | 79 (4) | 83 (3) | 82 (3) | 80 (4) | 46 | 109 |
| Accuracy (%) | 93.0 (1.6) | 93.5 (1.6) | 88.6 (2.2) | 89.7 (2.0) | 88.5 (1.8) | 90.1 (1.6) | 94.7 | 90.7 |
| Sensitivity (%) | 94.2 (2.1) | 95.4 (1.3) | 88.4 (4.0) | 92.7 (2.2) | 92.3 (2.0) | 90.2 (2.6) | 93.9 | 83.8 |
| Specificity (%) | 91.7 (2.6) | 91.5 (2.4) | 88.7 (3.1) | 86.7 (3.4) | 84.7 (3.8) | 90.0 (2.8) | 95.5 | 97.6 |
| Precision (%) | 92.0 (2.4) | 91.8 (2.2) | 88.8 (2.7) | 87.6 (2.8) | 85.9 (2.9) | 90.1 (2.5) | 95.4 | 97.2 |
| F1 Score (%) | 93.1 (1.5) | 93.6 (1.5) | 88.5 (2.3) | 90.0 (1.9) | 88.9 (1.6) | 90.1 (1.5) | 94.6 | 90.0 |
| MCC | 0.86 (0.05) | 0.87 (0.05) | 0.77 (0.05) | 0.79<br>(0.06) | 0.77 (0.06) | 0.80 (0.05) | 0.89 | 0.82 |

**Table 3.** Performance metrics for the risk assessment model using pairwise features on the 10 validation tests with six different classifier algorithms, final development testing, and deployed software blind test. Numbers correspond to individual’s classification average and standard deviations in parenthesis.

| Model | Risk assessment diagnosis development validation |  |  |  |  |  | Test | Blind Test |
| --- | --- | --- | --- | --- | --- | --- | --- | --- |
| Algorithms | ADA | GDB | RF | XRF | PLS | SVM | GDB | GDB |
| Vector length | 33 | 33 | 33 | 33 | 33 | 33 | 33 | 33 |
| # of Estimators | 260 (3) | 260 (3) | 68 (3) | 260 (3) |  |  | 256 | 256 |
| TN | 31 (3) | 31 (3) | 30 (3) | 30 (3) | 31 (3) | 31 (4) | 15 | 16 |
| FP | 6 (2) | 6 (3) | 7 (3) | 7 (3) | 6 (3) | 6 (4) | 7 | 5 |
| FN | 6 (2) | 6 (2) | 9 (2) | 9 (2) | 9 (2) | 8 (2) | 2 | 5 |
| TP | 33 (2) | 33 (2) | 30 (2) | 30 (2) | 30 (2) | 31 (2) | 21 | 34 |
| Accuracy (%) | 84.2 (4.1) | 84.5 (4.1) | 79.2 (4.2) | 79.4 (3.9) | 80.5 (4.2) | 81.2 (5.0) | 79.7 | 81.7 |
| Sensitivity (%) | 84.3 (5.5) | 85.3 (4.0) | 77.9 (5.4) | 77.1 (4.9) | 77.0 (5.0) | 78.7 (5.8) | 91.3 | 87.2 |
| Specificity (%) | 84.2 (5.8) | 83.7 (7.3) | 80.5 (8.2) | 81.7 (6.6) | 84.1 (8.2) | 83.7 (9.4) | 68.2 | 76.2 |
| Precision (%) | 84.4 (5.0) | 84.3 (5.9) | 80.5 (6.3) | 81.2 (5.6) | 83.5 (6.8) | 83.7 (8.2) | 74.2 | 78.5 |
| F1 Score (%) | 84.2 (4.2) | 84.7 (3.7) | 78.9 (3.9) | 78.9 (3.8) | 79.8 (4.0) | 80.8 (4.8) | 81.8 | 82.6 |
| MCC | 0.68 (0.11) | 0.68 (0.13) | 0.58<br>(0.12) | 0.58<br>(0.10) | 0.61<br>(0.13) | 0.63 (0.15) | 0.61 | 0.64 |

### Panel of discriminant metabolites for COVID-19 patients using untargeted metabolomics

Thirty ions were selected by the ML method and used for COVID-19 diagnosis using the introduced pairwise model (see ***Table 3*** for metrics) and further validated through mass spectrometric data. From those, we proposed 21 discriminant biomarkers for COVID-19 condition, divided into ten with positive (mean values higher for the positive group) and 11 with a negative contribution to the condition. Out of 21 molecules, eight belong to the glycerophospholipid class, three glycerolipids, three fatty acids, two cholesterol derivatives, one purine metabolite, one prostanoid, one plasmalogen, and two unknown peptides. The remaining ten molecules have not yet been identified, a common element of non-targeted metabolomics (14). Valid biomarkers and unknown features are available in ***Table 4***.

**Table 4.**
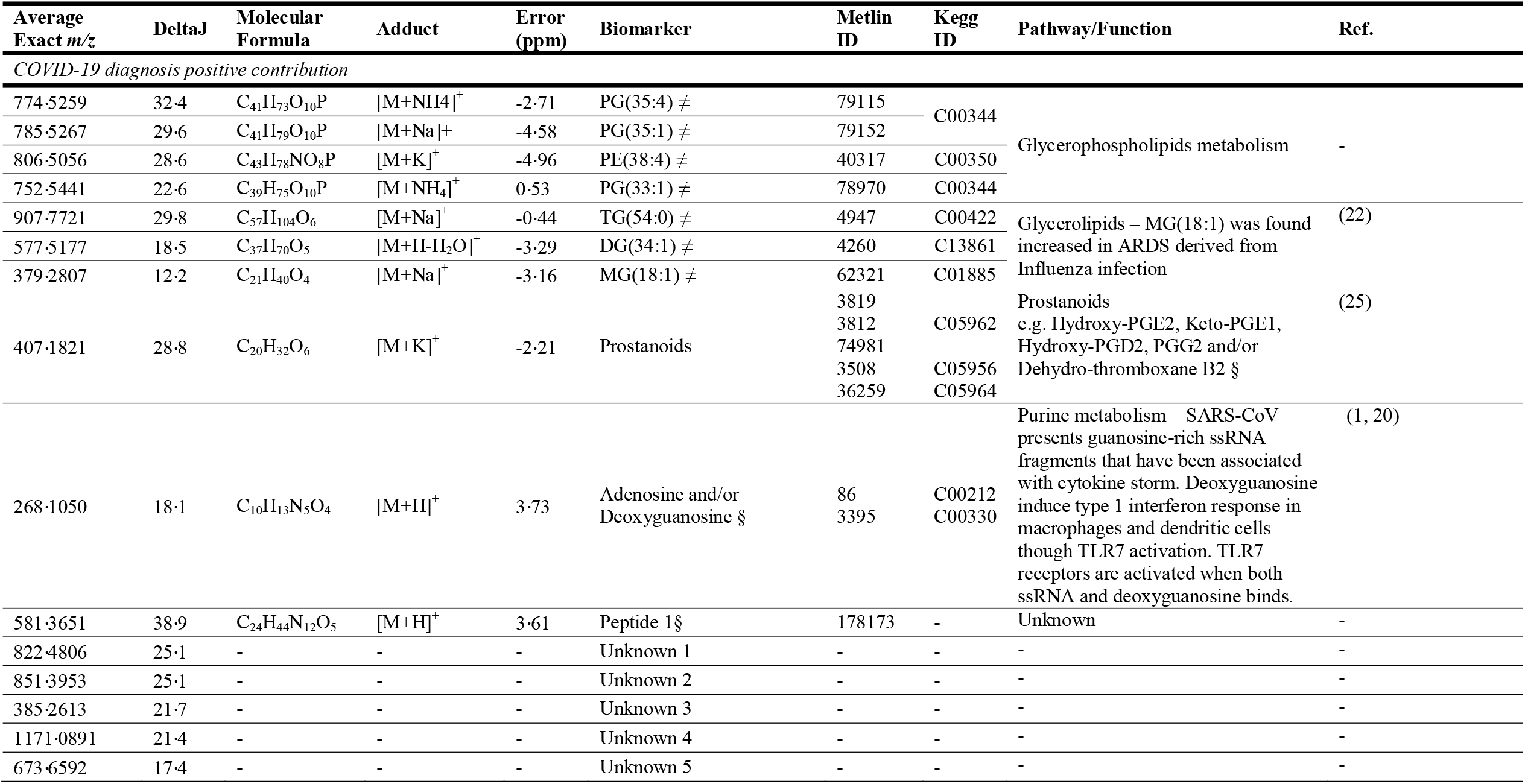
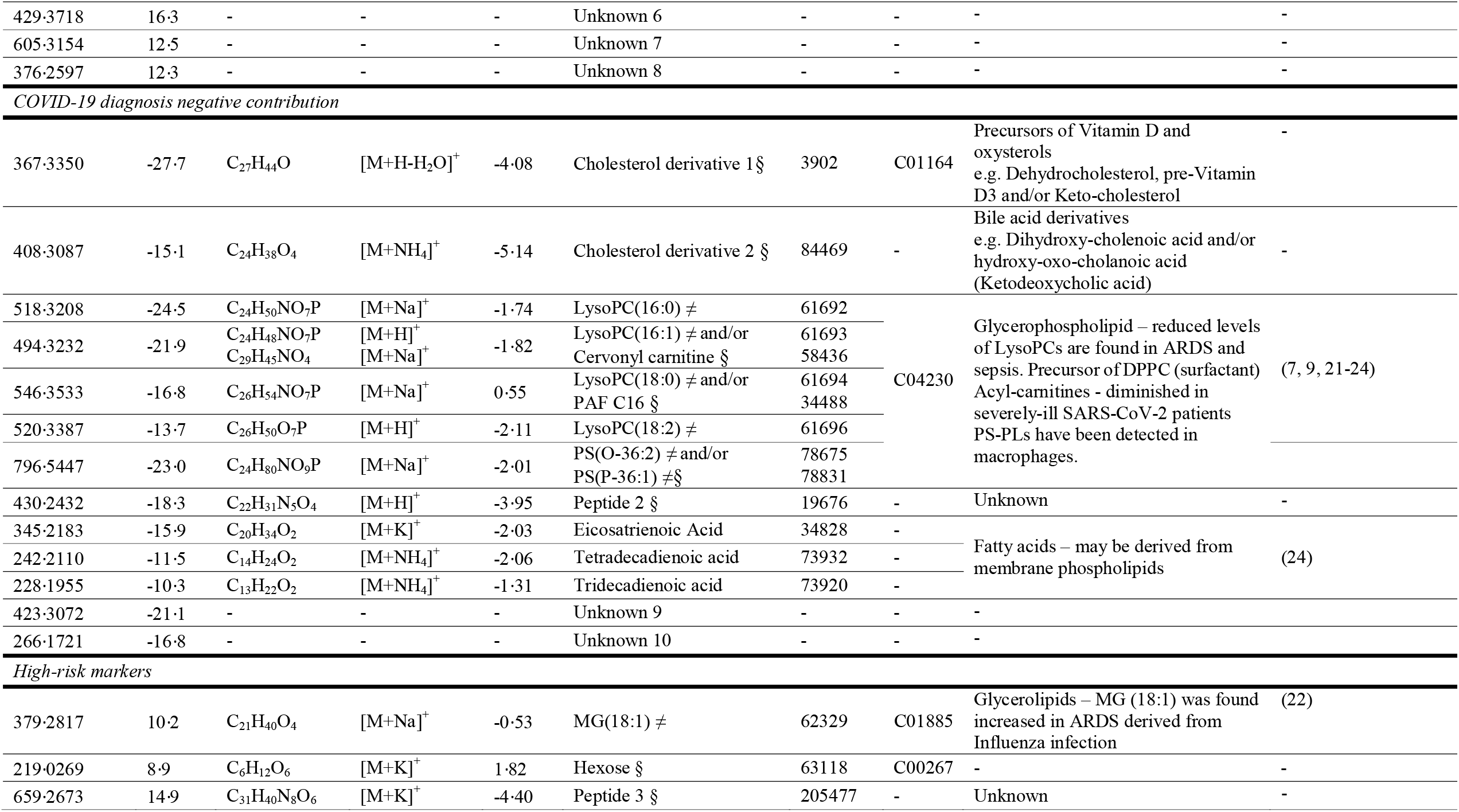
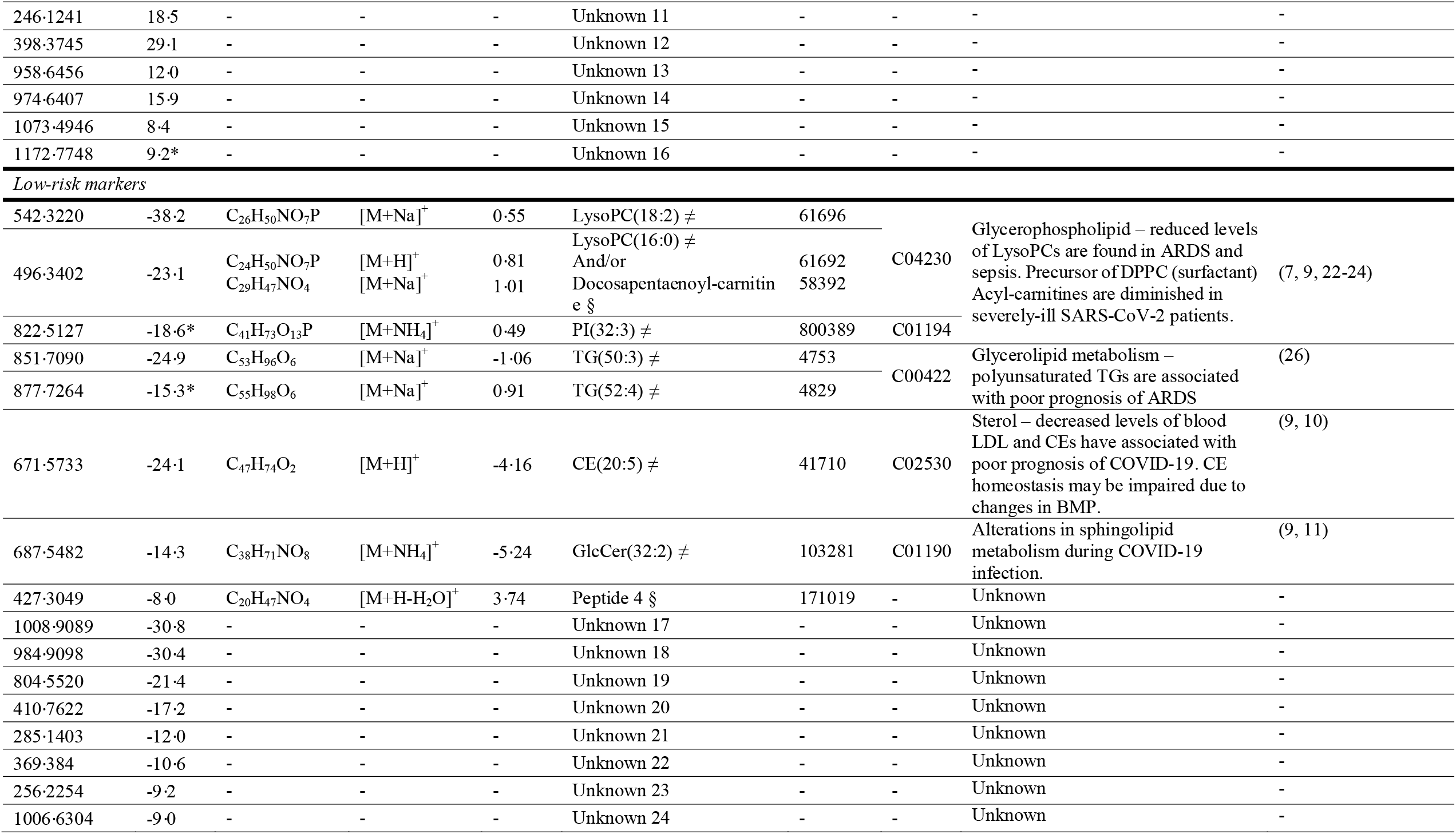
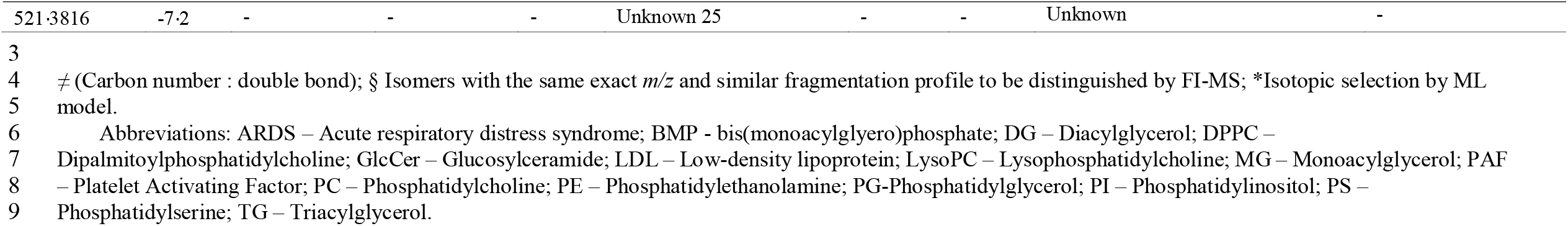
Proposed biomarkers to *m/z* discriminant features elected by Machine Learning algorithm group first by model contribution (COVID-19 diagnosis and risk assessment), followed by metabolic function and deltaJ.

For risk assessment, 26 ions were used to achieve the metrics displayed in ***Table 4***. Among them, nine biomarkers contributed to the COVID-19 higher risk condition and 17 biomarkers contributed to lower risk. The main findings shown in ***Table 4*** pointed to a relative reduction of certain species of lysophosphatidylcholine (LysoPC), phospholipids, cholesteryl ester (CE) and triacylglycerols (TG) in moderate/severe cases in comparison to patients with mild symptoms (***Figure 2a***). In ***Table 4*** the biomarkers were first grouped by type of contribution, followed by metabolic class/function and importance reflected through ⍰J metric. A representation of biomarkers class and ΔJ metrics are displayed in ***Figure 2a***.

## DISCUSSION

### MS-ML elected biomarkers and COVID-19 pathophysiology

The use of AI-explained algorithms allowed us to create reliable models that facilitate decision-making in clinics and the investigation of the pathophysiological meaning of the distinct biomarker’s levels. Viral recognition is an essential step for initial host immune response, and the rapid course and cytokine storm associated with SARS-CoV infection may be involved with the guanosine- and uridine-rich (GU) single-strand RNA potential role as PAMP (pathogen-associated molecular patterns) (1). Deoxyguanosine [268·1050, [M+H]^+^), a metabolite from purine metabolism (Kegg hsa00230), triggers the enhanced signalling of TLR7 in the presence of ssRNA, inducing cytokine secretion in macrophages (20). Therefore, further investigations are required to understand the potential role of deoxyguanosine in SARS-CoV-2 immune hyperactivation and pathology.

The main lipidic findings pointed to a remodelling of glycerophospholipid metabolism. We identified enhanced presence of phosphatidylglycerol (PG) [PG(35:4), PG(35:1), PG(33.1)] and phosphatidylethanolamine (PE) [PE(38:4)], and a diminishment of lysophosphatidylcholines (LysoPC) [LysoPC(16:0), LysoPC(16:1), LysoPC(18:0), LysoPC(18:2)] and phospatidylserine plasmalogens (PS-PL) (21) [PS(O-36:2) and/or PS(P-36:1)] in COVID-19 positive patients, as illustrated in ***Figure 2a*** by glycerophospholipid pathway recurrence.

LysoPCs [LysoPC(16:0) and LysoPC(18:2)] were also found as negative contributors in plasma samples from patients who required hospitalization (moderate and severe cases). Cell responses to various stimuli may be mediated by phospholipids, which actively participates in inflammation processes. The relative intensities decrease of Lysophosphatidylcholines in positive and, and some of them, in moderate to severely-ill patients, are in accordance with recent studies of metabolic changes in acute respiratory distress syndrome (ARDS) and sepsis (22, 23), important characteristics of COVID-19 severity (2, 7).

LysoPC is formed through the cleavage of PC mediated by phospholipase A_2_, (PLA_2_), whose modulation has a crucial role in inflammation processes (see LysoPCs’ related pathways in ***Figure 2b***). PLA_2_ up-regulation promotes fatty acids formation, precursors of eicosanoids, and LysoPCs (24). Data show that SARS-CoV nucleocapsid protein stimulates the expression of Ciclooxygenase-2 (COX-2), an essential enzyme in the catalyses of prostanoids production from fatty acids, as those found at *m/z* 407.1821 in positive group (25). Although we identified an ion correlated to eicosanoid biosynthesis that indicates PLA_2_ and COX-2 activity in positive patients, LysoPCs were relatively decreased in this group. The availability of LysoPCs is also finely regulated by the acyltransferase activity of LCAT (Lysophosphatidylcholine Acyltransferase 1), which may promote the restoration of PCs via Lands cycle. The most abundant lipid species found in alveolar surfactant formed by LCAT1 activity over LysoPC is Dipalmitoylphosphatidylcholine (DPPC, PC(16:0/16:0)). This molecule corresponds to 70-80% of surfactant lipid composition, and the dysregulation of surfactant film is directly related to lung injury and ARDS (24). Since DPPC formation is dependent on the availability of lipid substrates and the Lands cycle functioning, interferences in this process may disturb LysoPC availability. In a metabolomic study, Ferrarini et al (2017) described a decrease in LysoPC species and increased MG(18:1) [*m/z* 379.2807] in serum of patients with ARDS derived from Influenza infection and sepsis, reinforcing our findings (22).

Moreover, COVID-19 pathophysiology seems to impair cholesterol homeostasis (9, 10). We found cholesteryl ester (CE) associated with mild symptoms, which was similarly reported by Song et. al (2020). They demonstrated the correlation between CE abundance and BMP(38:5), a lipid that influences cellular exportation of cholesterol from endosomes. During recovering progression, it was found an increased alveolar macrophages BMP with enhanced CEs (9). Cholesterol and LDL (low-density lipoprotein) lowering was also observed in clinical practice associated with COVID-19 poor prognosis (10), such as triacylglycerol in ARDS (26).

Herein, based on the proposed *m/z* ions we discriminated COVID-19 patients using a diagnostic and risk assessment classifier generated from a MS-ML combination. Although the proposed biomarkers correlates COVID-19 pathophysiology to the mathematical process, a more comprehensive biomarker evaluation is needed to better understand their contribution to COVID-19, and identify the unknowns.

### Use of untargeted metabolomics and ML for automated COVID-19 diagnosis and risk assessment

The combination of artificial intelligence algorithms for biomarker mining in complex data is a common approach for problem-solving and implementing new technologies in health sciences. The use of machine learning as a mean for the discrimination of diseases from mass spectrometric data aims to develop diagnostic and prognostic biomarkers, treatment targets and patient management systems (13).

Our methodology introduced the pairwise *m/z* analysis, an essential advance in untargeted metabolomics application. By combining different *m/z*, this approach supports the spectra acquired by different mass spectrometers, including the robust use of flow-injection mass spectrometry (FI-MS), in an effort to overcome the ion competition effect (27).

The model optimization with pairwise features can be easily transferred to an independent diagnosis platform. Given that the process key is available from biological sample “ion-fishing”, this approach does not require chromatography and biomarkers quantitation for independent diagnosis. Moreover, the proposed MS-ML platform for COVID-19 presented reliable qualitative results, with specificity of 97·6% and sensitivity of 83·8% (in a blind test data), similar or even better in performance when compared to available serology (5) and RT-PCR methods (6). Our analysis also brings molecular information about disease pathophysiology that may aid in prognostic markers and treatment targets for COVID-19. Overall, our test aggregates, in one solution, an alternative for populational COVID-19 screening and guidance for public health efforts through risk classification. The same approach may be applied to other diseases involved with patient management during the pandemic and contribute to the COVID-19 MS Coalition’s collective effort (15) by consolidating the combination of mass spectrometry and artificial intelligence in a real-world setting.

## Data Availability

The data generated in this study will be anonymized to attend patient's privacy restrictions and will be available from corresponding authors upon request after publication.

## ACKNOWLEDGMENTS

The authors would like to thank the network involved in sample collection, clinical and diagnosis support, and Thermo Scientific and LADETEC (UFRJ) for technology support.

## AUTHOR CONTRIBUTIONS

R.S.F., J.C.N., T.F.D., A.J.B., E.C.S., L.O.R., W.J.F., M.W.P.J., M.V.G.L., G.C.M., W.M.M., F.F.A.V., D.B.S., V.S.S., J.D., and R.R.C were involved with study design and ethics approval. R.S.F., T.F.D., A.J.B., R.S., E.C.S., A.E., L.O.R., W.J.F., N.D., L.A.S, J.C.C.A., M.V.G.L., G.C.M., W.M.M., F.F.A.V., R.L.A.N., D.B.S. and V.S.S. contributed to patient data acquisition and analysis, clinical support and network feasibility. L.C.N., J.D., A.R.R. and R.R.C., conceived and developed the MS-ML method. J.D., E.N.BB., G.M.S., A.N.O., D.N.O. performed mass spectrometry experiments and data interpretation. J.D., L.C.N., R.F.S. prepared tables, figures and wrote the manuscript. E.N.B.B., G.M.S., A.N.O., D.N.O., F.T.M.C., N.D., W.F.J., L.O.R., D.B.S., F.F.A.V., W.M.M., G.C.M., J.C.N., A.R.R. and R.R.C. revised the manuscript. RRC idealized the project and managed the research group. All authors read and approved the manuscript.

